# Towards personalized nicotinamide mononucleotide (NMN) supplementation: nicotinamide adenine dinucleotide (NAD) concentration

**DOI:** 10.1101/2024.02.19.24303025

**Authors:** Ajla Hodzic Kuerec, Weilan Wang, Lin Yi, Rongsheng Tao, Zhigang Lin, Aditi Vaidya, Sohal Pendse, Sornaraja Thasma, Niranjan Andhalkar, Ganesh Avhad, Vidyadhar Kumbhar, Andrea B. Maier

**Author notes:** These two authors contributed equally (Ajla Hodzic Kuerec, Weilan Wang). corresponding author: Professor Andrea Maier.

## Abstract

Nicotinamide mononucleotide (NMN) is a precursor of nicotinamide adenine dinucleotide (NAD), which declines with age. Supplementation of NMN has been shown to improve blood NAD concentration. However, the optimal NMN dose remains unclear. This is a post-hoc analysis of a double-blinded clinical trial involving 80 generally healthy adults aged 40 to 65 years. The participants received a placebo or daily 300 mg, 600 mg, or 900 mg NMN for 60 days. Blood NAD concentration, blood biological age, homeostatic model assessment for insulin resistance, 6-minute walk test, and 36-item short-form survey (SF-36) were measured at baseline and after supplement. A significant dose-dependent increase in NAD concentration change (NAD_Δ_) was observed following NMN supplementation, with a large coefficient of variation (29.2-113.3%) within group. The increase in NAD_Δ_ was associated with an improvement in the walking distance of 6-minute walk test and the SF-36 score. The median effect dose of NAD_Δ_ for the 6-minute walk test and SF-36 score was 15.7 nmol/L (95% CI: 10.9-20.5 nmol/L) and 13.5 nmol/L (95% CI; 10.5-16.5 nmol/L), respectively. Because of the high interindividual variability of the NAD_Δ_ after NMN supplementation, monitoring NAD concentration can provide valuable insights for tailoring personalized dosage regimens and optimizing NMN utilization.

**Highlights:**

- Large interindividual variability in blood NAD response to NMN supplement was observed.
- The median effective dose of blood NAD concentration improvement to achieve clinically significant improvement in functional outcome and quality of life was around 15 nmol/L.
- A close monitoring of NAD concentration change and personalized regimen of NMN supplements is needed.

## 1. Introduction

Nicotinamide mononucleotide (NMN) is a bioactive nucleotide formed by the reaction between a nucleoside containing ribose, nicotinamide (NAM), and a phosphate group (Poddar et al., 2019). It is a precursor of nicotinamide adenine dinucleotide (NAD), a crucial cofactor for enzymes involved in major biological processes such as cellular redox regulation and metabolism, and DNA repair (Fouquerel and Sobol, 2014; Koju et al., 2022; Williamson et al., 1967). Blood NAD concentration declines with chronological age in animals and humans (Chaleckis et al., 2016; Clement et al., 2019; Frederick et al., 2016; Gomes et al., 2013; Mouchiroud et al., 2013) due to the declined de novo synthesis of NAD and the hyperactivity of NAD-consuming enzymes (McReynolds et al., 2021). Preventing the depletion of NAD concentration in *in-vitro* studies had neuroprotective (Liu et al., 2008), cardioprotective effects (Pillai et al., 2005), and promoted DNA repair (Wilk et al., 2020).

Dietary supplementation with NMN and other NAD precursors has been shown to increase the level of NAD concentration in the blood of middle-aged and older individuals (Yi et al., 2023; K et al., 2022; Karol M Pencina et al., 2023; Huang, 2022). While inter-individual variability in the increase of blood NAD after NMN supplementation has been observed in previous studies (K et al., 2022; Karol M Pencina et al., 2023), there are no studies reporting the relationship between change in blood NAD concentration (NAD_Δ_) following NMN supplementation and aging-related clinical outcomes.

This study reports the determinants of blood NAD basal level and NAD_Δ_, the relationship between the blood NAD concentration and the change in blood biological age (blood biological age_Δ_), change in homeostatic model assessment for insulin resistance (HOMA-IR_Δ_), change in 6-minute walk test (6-minute walk test_Δ_), change in 36-item short form survey scores (SF-36_Δ_), and the variance and median effective dose (ED_50_) of NAD_Δ_ after 30 or 60 days of NMN supplementation in healthy middle-aged individuals.

## 2. Material and method

### 2.1. Study design

The study design of this double-blinded, randomized clinical trial was reported elsewhere (Yi et al., 2023), The trial was conducted in Lotus Healthcare and Aesthetics Clinic and Sunad Ayurved in Pune, India, and monitored by a clinical research organization (CRO) in Pune, ProRelix Services LLP. Briefly, generally healthy 40 to 65 years old individuals with a body mass index between (BMI) 18.5 to 35 kg/m2 were included in this study. Participants were randomized into four groups receiving placebo or daily NMN supplements of 300 mg, 600 mg, and 900 mg, respectively, for 60 days. Age, sex, BMI, blood biological age (calculated by Aging.AI 3.0) (Putin et al., 2016) and HOMA-IR were recorded at baseline and day 60. In addition, blood NAD concentration, 6-minute walk test, and 36-item short-form survey (SF- 36) scores were measured at baseline, 30 days, and 60 days. All participants provided written informed consent. The ethical approval was obtained from the Royal Ethics Committee, Pune, India.

### 2.2. Data Analysis

The normality of continuous variables was tested by the Shapiro-Wilk test. Data were presented as means and standard deviations (SD) or median and inter-quantile range (IQR) according to normality for continuous variables or numbers and percentages (%) for categorical variables. NAD_Δ_ was defined as the difference in blood NAD concentration at day 30 or day 60 from the NAD concentration at baseline. The inter-group variance of NAD_Δ_ was assessed by the coefficient of variance (CV). The difference in blood NAD concentration after supplementation was investigated by the Kruskal-Wallis test and the group-to-group difference was investigated by Dunn’s post-hoc test. Normalization for skewed data was conducted using square root transformation. The association between chronological age, blood biological age, sex, body mass index (BMI), and homeostatic model assessment for insulin resistance (HOMA-IR) and blood NAD baseline concentration or NAD_Δ_ at day 30 or day 60 were investigated by generalized linear regression using Gaussian method.

Generalized linear regression was also used to investigate the association between NAD_Δ_ and changes in blood biological age, HOMA-IR, and changes in the 6-minute walk test and SF-36 scores. The outlier for the generalized linear regression was defined as a data point where studentized residual over ±2. Outliers were excluded from the generalized linear regression analysis. Outcomes from generalized linear regression are shown as regression coefficient and 95% confidence interval (95% CI).

The median effective dose (ED_50_) is a dose required to achieve a targeted effect in 50% of the population receiving the dose. In this study, ED_50_ of NAD_Δ_ after a 60-day NMN supplementation was calculated to determine a target NAD concentration improvement needed to achieve significant clinical improvement in a significant proportion of the population. Significant clinical improvement for the 6-minute walk test was defined as an increase of at least 30 meters from the baseline outcome (Bohannon and Crouch, 2017) and for the SF-36 score as an increase of at least ten points from the baseline score (Bjorner et al., 2007). To ensure a wide range of NAD_Δ_ and a sufficient sample size in each group, the participants were divided into 20 groups (four individuals in each group) based on their NAD_Δ_, to simulate having 20 dose groups in pharmacological studies. The NAD_Δ_ of each group was re-calculated as the mean value of the NAD_Δ_ of the four individuals in the group. A four-parameter log-logistic model was used to assess the dose-response effect, where dose referred to NAD_Δ_, and response referred to achieving significant clinical improvement or not. The 95% confidence interval of ED_50_ was obtained using the delta method, a method to calculate the approximated standard error for results generated from the function.

Data analysis was performed on R (version 4.2.1). ED_50_ was calculated using the “drc” package. A p-value of less than 0.05 was considered statistically significant.

## 3. Results

The demographic characteristics and responses are shown in Table 1. The number of male participants was eight in the placebo group, ten in the 300 mg group, six in the 600 mg group, and nine in the 900 mg group. All participants showed good compliance, ranging from 75% to 100%.

**Table 1.**
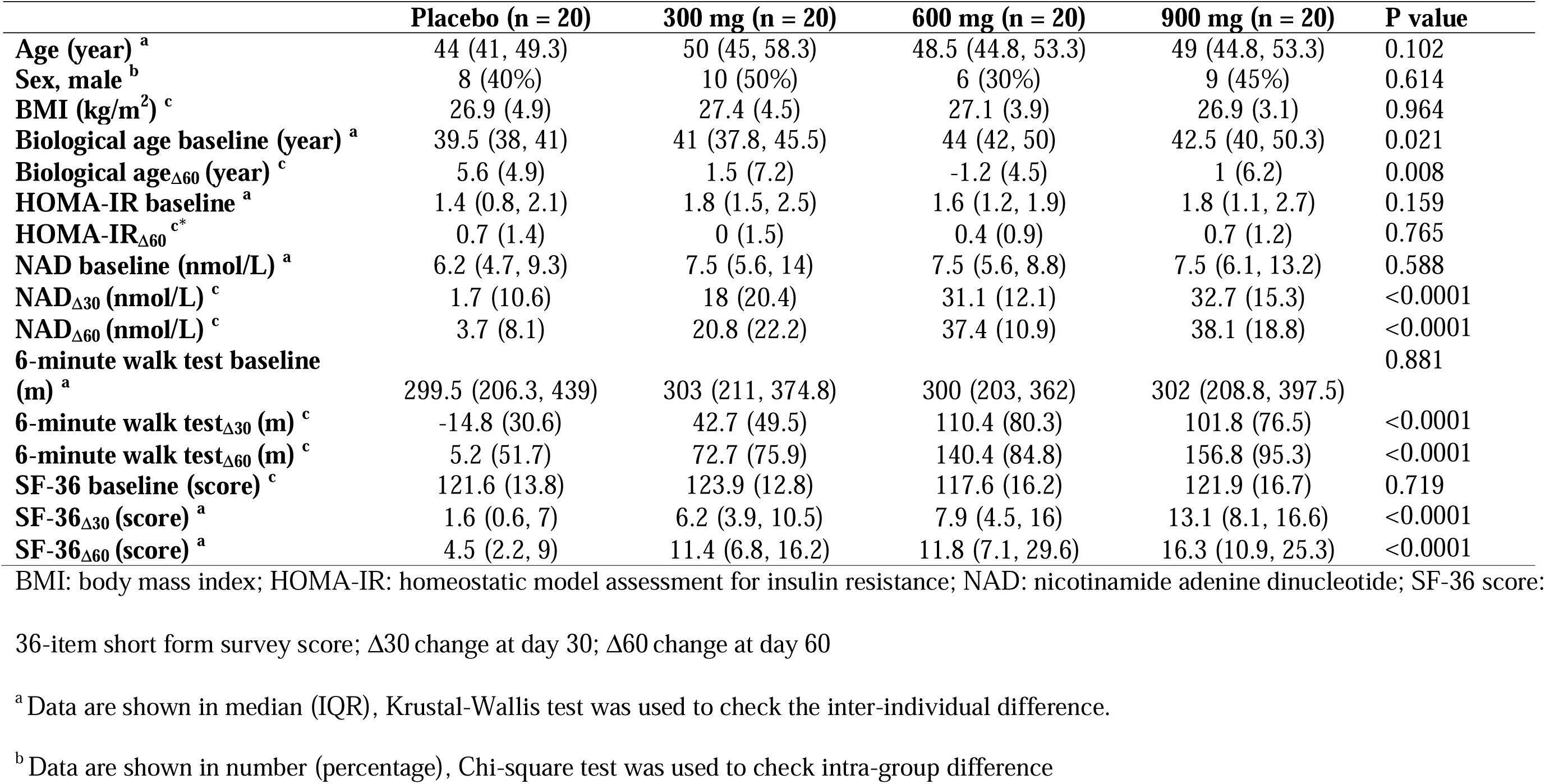
Participant characteristics at baseline and at day 30 and day 60.

The NAD_Δ_ from baseline to day 30 and day 60 is shown in Figure 1. The NAD_Δ_ at both day 30 and day 60 increased dose-dependently (p<0.001), while the SD within the group was large, the CV of blood NAD concentration after supplementation ranging from 220% to 614% in the placebo group, and from 29.2% to 113.3% in intervention groups. The NAD_Δ_ was dose-dependent with no significant difference between the 600 mg group and the 900 mg NMN group.

**Figure 1.**
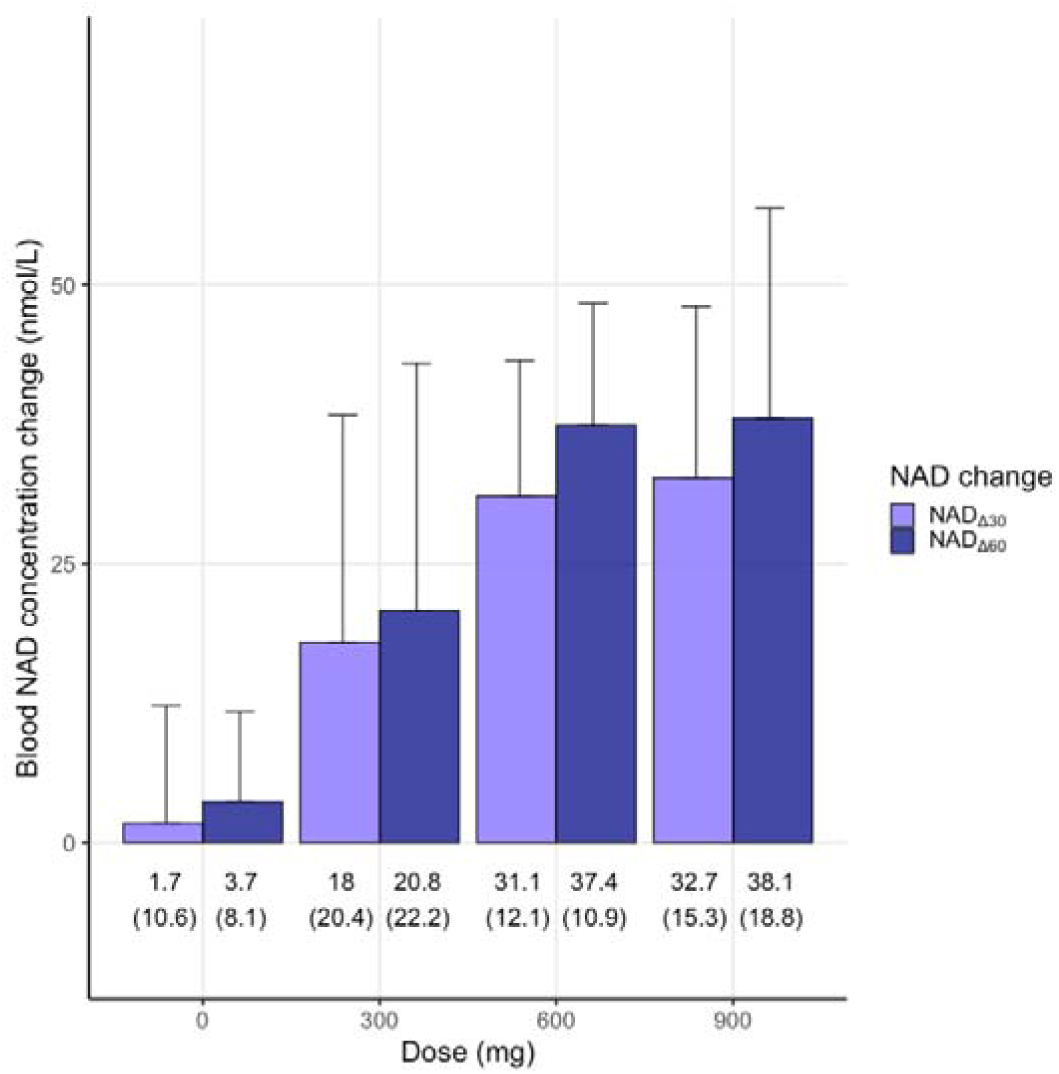
Blood NAD concentration change in each dose group. NAD: nicotinamide adenine dinucleotide; Δ30: change at day 30; Δ60: change at day 60 Data are shown in mean (SD). *colour should be used

No significant association was found between blood NAD baseline concentration or NAD_Δ_ at day 30 or day 60 and chronological age or blood biological age, sex, and BMI. A higher baseline HOMA-IR ratio was associated with higher baseline NAD concentration but not with NAD_Δ_ (Table 2).

**Table 2.**
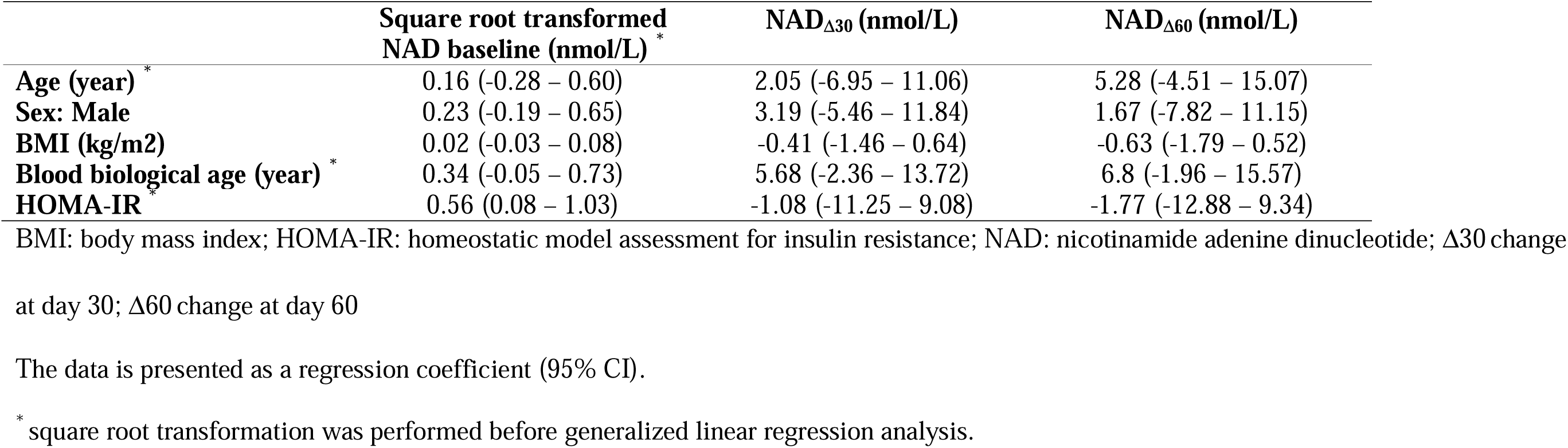
The association of participant characteristics at baseline with NAD concentration levels Square root transformed NAD baseline (nmol/L) *.

No significant association was found between blood baseline NAD concentration and 6-minute walk NAD_Δ_, SF-36 score_Δ_, blood biological age_Δ_, and HOMA-IR_Δ_ (Table 3). Higher NAD_Δ_ was significantly associated with better improvement in 6-minute walk_Δ_ and SF-36 score_Δ_ at both day 30 and day 60. Each 1 nmol/L increase in blood NAD concentration at day 30 was associated with 1.37 meters more in a 6-minute walk test. Each 1 nmol/L increase in blood NAD concentration was also associated with 0.02 more in the square root of SF-36 scores. For example, 5 nmol/L blood NAD concentration increase was associated with 5.76 (5*0.02+2.30 intercept)^2 higher SF-36 score at day 30. The improvements associated with higher NAD_Δ_ were larger in measurements taken after 60 days of supplementation.

**Table 3.** The association between NAD baseline level or NAD change and aging-related clinical outcomes.

| | 6-minute walk testΔ<br>(meter) | Square root transformed<br>SF-36 scoreΔ<sup>*</sup> | Biological ageΔ (year) | HOMA-IRΔ |
| --- | --- | --- | --- | --- |
| **NAD baseline (nmol/L)** <sup>a*</sup> | 1.27 (-20.16 – 22.69) | 0.27 (-0.08 – 0.61) | -0.79 (-2.07 – 0.5) | 0.18 (-0.17 – 0.52) |
| **NAD$\Delta_{30}$ (nmol/L)** <sup>b</sup> | 1.36 (0.67 – 2.05) | 0.02 (0.01 – 0.03) | NA | NA |
| **NAD$\Delta_{60}$ (nmol/L)** <sup>a</sup> | 2.43 (1.43 – 3.42) | 0.02 (0.01 – 0.04) | -0.02 (-0.08 – 0.03) | 0 (-0.01 – 0.01) |

HOMA-IR: homeostatic model assessment for insulin resistance; NA: not applicable; NAD: nicotinamide adenine dinucleotide; NAD $\Delta_{30}$ :
nicotinamide adenine dinucleotide at day 30; NAD $\Delta_{60}$ : nicotinamide adenine dinucleotide at day 60; SF-36 score: 36-item short form survey
score; $\Delta$ change
The data is presented as a regression coefficient (95% CI).
<sup>a</sup> The linear regression was calculated by NAD baseline or NAD $\Delta$ at day 60 and 6-minute walk test $\Delta$ at day 60 or SF-36 $\Delta$ at day 60
<sup>b</sup> The linear regression was calculated by NAD $\Delta$ at day 30 and 6-minute walk test $\Delta$ at day 30 or SF-36 $\Delta$ at day 30.
<sup>\*</sup> square root transformation was performed before generalized linear regression analysis

Each 1 unit increase of the NAD level on day 60 was associated with 2.42 meters more in the 6-minute walk test and 0.02 more in the square root of SF-36 scores. Thus, 5 nmol/L NAD concentration increase is associated with 8.82 (5*0.02+2.87 intercept)^2 higher SF-36 score on day 60. No significant association was found between NAD_Δ_ and blood biological age_Δ_ or HOMA-IR_Δ_.

After the 60-day supplementation, a total of 49 and 40 participants had a significant improvement in the 6-minute walk test and SF-36 score, respectively. The ED_50_ of NAD_Δ_ at day 60 for the clinically significant improvement of the 6-minute walk test and SF-36 score was 15.65 nmol/L (95% CI: 10.87-20.45 nmol/L) and 13.51 nmol/L (95% CI; 10.54-16.50 nmol/L) respectively. The dose-response curves of the NAD_Δ_ and the percentage of respondents having a 6-minute walk test_Δ_ at day 60 ≥30 meters or SF-36_Δ_ at day 60 ≥10 scores are shown in Figure 2.

**Figure 2.**
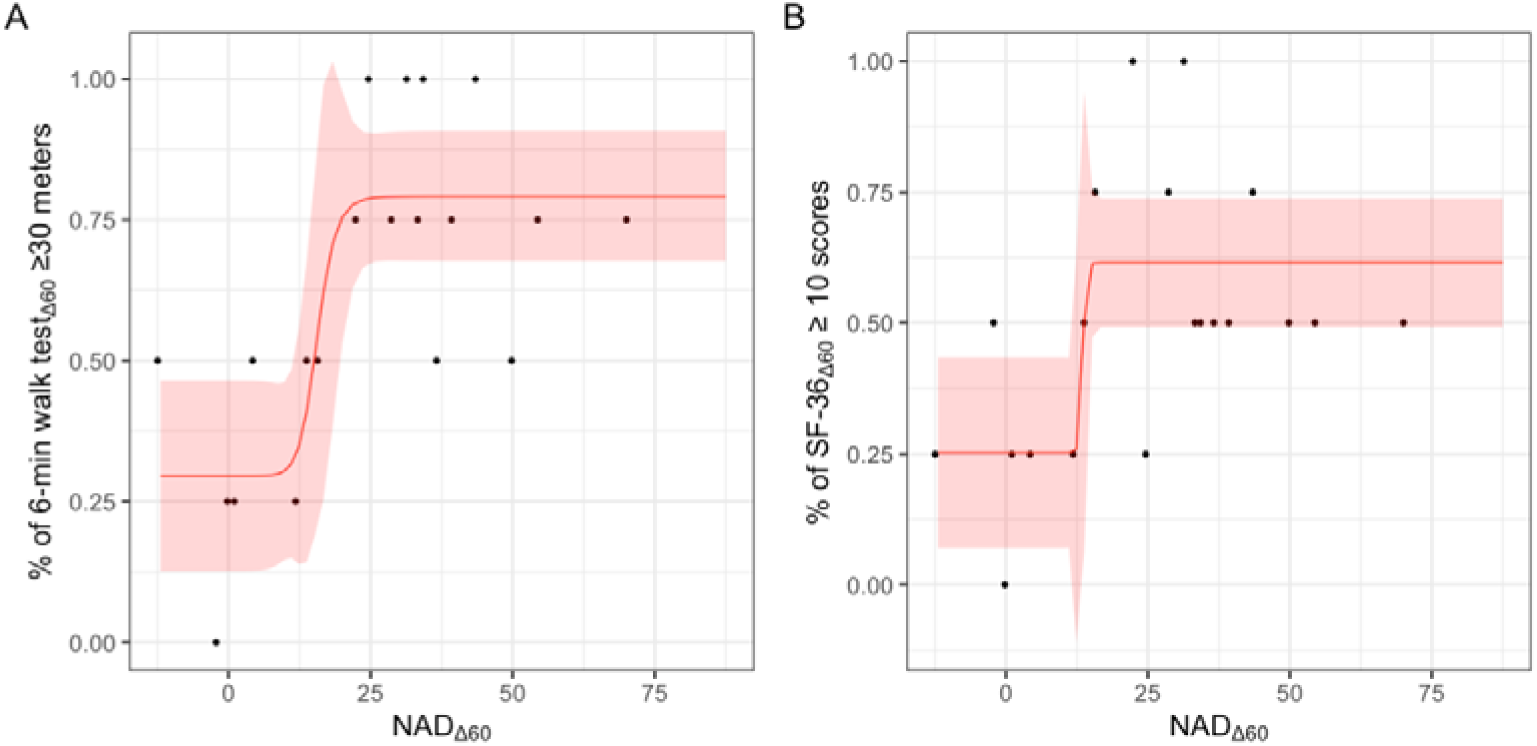
The dose-response effect curve of blood NAD concentration changes to clinically significant improvement in (A) 6-minute walk test_Δ_and (B) SF-36 score_Δ_ from baseline to 60 days. *colour should be used

## 4. Discussion

This post-hoc analysis of a randomized, double-blinded, controlled clinical trial of NMN supplementation indicates that blood NAD concentration changes are dose-dependent but exhibit high interindividual variability. Only HOMA-IR was significantly associated with blood baseline NAD. Higher NAD_Δ_ were associated with improvements in 6-minute walk test distance and SF-36 scores and NAD_Δ_ ≈ 15 nmol/L produced a clinically significant improvement in these outcomes.

In the efficacy and safety analysis of the same study, significant improvement in the 6-minute walk test and SF-36 score from baseline were reported in the 600 mg group and 900 mg group, while no significant improvement in the 6-minute walk test was seen in the 300 mg group (Yi et al., 2023). The lack of association in the 300 mg group may be due to the large variation in blood NAD response to NMN supplementation, especially in a lower dose. In this post hoc analysis, significant associations were found between both the 6-minute walk test and the SF-36 score and NAD_Δ_, indicating the efficacy of NMN supplementation should not only be assessed through the dosage of NMN administrated but also on the individual’s actual NAD response.

Whole blood NAD concentrations after supplementation with NMN are highly variable in healthy middle-aged individuals (K et al., 2022; Karol M Pencina et al., 2023; Yi et al., 2023), no significant associations were found between basal NAD concentration levels and chronological age and gender, which is in concordance with studies in healthy individuals (Yang et al., 2022) and in adults with hypertension, diabetes mellitus, and hyperlipidemia (Breton et al., 2020). However, other studies reported that men had significantly higher basal blood NAD concentrations than women (Breton et al., 2020; Yang et al., 2022). These results indicate that the association between NAD concentration levels and sex and underlying mechanisms should be explored in future studies.

Similar to our results, among healthy Japanese individuals aged 20-65 years, chronological age and BMI were not significantly associated with NAD_Δ_ after supplementation of 250 mg NMM for 12 weeks (K et al., 2022). The gut microbiome could potentially influence the availability of NMN, as previous studies found a bidirectional cycling of NAD precursors between the human bloodstream and gut microbiota (Chellappa et al., 2022). Mutations on specific genes such as GLUL lead to NAD+ synthesis rate change (Hu et al., 2015). However, the association between gut microbiome species and abundance or gene mutation and NAD synthesis after NMN supplementation is still unknown. Mutations on specific genes such as GLUL lead to lower NAD+ synthesis rate (Hu et al., 2015). However, the association between gut microbiome species and abundance, gene mutation, and NAD synthesis after NMN supplementation is still unknown. Other possible factors that could affect the variability in NAD_Δ_ are the difference in the bioavailability of NMN, concentrations, and activity of the NAD+ salvage enzymes such as nicotinamide phosphoribosyl transferase (NAMPT), which utilize NMN for the production of NAD (Akan et al., 2022; de Guia et al., 2019) and the consumption of NAD (Almangush et al., 2020). The NAMPT abundance in musculoskeletal system is associated with higher physical activity, indicating that people with more sedentary lifestyle may have a lower NAD_Δ_ (de Guia et al., 2019). Furthermore, the overexpression of cluster of differentiation 38 (CD38), whose major enzymatic activity lies in the hydrolysis of NAD, is associated with lower cellular NAD levels in multiple mammalian tissues (Aksoy et al., 2006). However, intra-cellular changes were not explored in this clinical trial.

The 6-minute walk test is used for the assessment of the progression of functional exercise capacity in clinical intervention studies (Rubenstein et al., 2000) with high reliability of the test in healthy older individuals (Rikli and Jones, 1998). The increase in the blood NAD concentration was associated with an improvement in the 6-minute walking distance, reflecting better physical performance. This is the first study that describes the relationship between blood NAD concertation and a functional outcome parameter. SF-36 score is a validated tool for the assessment of health-related quality of life. It consists of eight domains with separate scoring for each domain: physical functioning, role physical, bodily pain, general health, vitality, social functioning, role emotional, and mental health (Rubenstein et al., 2000). The increase in the blood NAD concentration was associated with a higher overall SF-36 score. Future studies should explore the relationship of blood NAD concentration change with each domain separately (Rubenstein et al., 2000). The increase in NAD+ levels resulting from NMN supplementation may lead to improvements in the 6-minute walk by enhancing mitochondrial function with increased ATP production and subsequently providing higher energy availability and endurance during physical activity (Ji and Yeo, 2022). Furthermore, enhanced NAD concentration is associated with better cardiovascular function by promoting vasodilation, increasing blood flow, and enhancing overall cardiovascular performance (Das et al., 2018; Zhang et al., 2016).

Biological ageing takes place as a result of the gradual accumulation of pathophysiological changes unrelated to diseases (Jazwinski and Kim, 2019; Wu et al., 2021). A biological age clock is used to predict a person’s lifespan and health span (Jylhävä et al., 2017). In this study, the biological age was calculated by the Aging.AI 3.0 from 19 blood biomarkers including complete blood count, lipids, and minerals (Mamoshina et al., 2018; Putin et al., 2016).

Although significant improvements were seen in physical performance and general health, no significant association between NAD_Δ_ and blood biological age was found in our study, which may be due to a relatively small sample size or the lack of effect. A 60-day supplementation may not be able to alter the blood biomarkers to a noticeable level. Further studies with a larger cohort should explore the relationship between blood NAD concentration levels and biological age for a longer duration.

Although there was no association between HOMA-IR and the change in blood NAD concentrations in our study, preclinical studies report that nicotinamide nucleotide adenylyltransferase 3 (Nmnat3) (an enzyme that degrades NAD and generates NAM as a by- product (Hikosaka et al., 2014)) overexpression in mice efficiently increased NAD concentration levels in various tissues and prevented diet-induced and aging-associated insulin resistance (Gulshan et al., 2018).

NMN supplementation was reported to be safe, with adverse events when present, typically being mild or moderate and manageable in clinical trials. (Huang, 2022; Yi et al., 2023) No significant adverse effect was observed after supplementation of daily 250 mg NMN for 24 weeks in a group of men with diabetes and impaired physical performance aged 65 years or above (Barker et al., 2022). It was also shown to be safe with high doses in a short duration. No severe adverse event was reported after a daily supplement of 2000 mg β-NMN for 4 weeks among middle-aged and older adults (Pencina et al., 2023, 2). However, further studies should assess the safety profile of NMN when given for a longer duration.

An increase in blood NAD concertation due to NMN supplementation was observed during the intervention periods in human trials (Huang, 2022; Pencina et al., 2023; Yi et al., 2023) however, clinical trials that include a follow-up period after the completion of the intervention period are lacking to determine the long-term effect of NMN interventions on NAD concentrations and its association with clinical outcomes, which are needed to determine the optimal treatment duration. In a study where two MIB-626 tablets, each containing 1000 mg of β-NMN, were administrated two times a day for 28 days to middle-aged overweight or obese participants, blood NAD concentration increased significantly, while NAD levels returned to the baseline 28 days after the last dose. However, the association between clinical outcomes and the NAD level during the intervention and follow-up of this study was not assessed (Pencina et al., 2023, 2). Therefore, the assessment of the long-term impact of NMN supplementation is warranted in future clinical trials.

Furthermore, a cross-sectional study in China reported that individuals with blood NAD concentration ≥36.4 nmol/L had a three times higher risk of having metabolism diseases than those with <29.4 nmol/L (Liu et al., 2023). A significant positive association between blood baseline NAD concentration and higher HOMA-IR was also found in this trial. Higher blood NAD concentrations might, therefore not perse be beneficial. This is the first study that reports the ED_50_ of NAD_Δ_ for the improvement in the 6-minute walk test and SF-36 score, which could serve as an indication for NMN titration in clinical practice. By closely monitoring changes in blood NAD concentration, a personalized dosage can be administered, aiming to achieve optimal effects at the individual level. It is important to note that no established guideline for an optimal therapeutic window of blood NAD concentration exists, thus closer monitoring of blood NAD for a longer follow-up duration is needed in future studies. All results have to be treated with caution due to the relatively small sample size.

Factors that could influence baseline NAD concentrations should be taken into account in future larger trials.

## 5. Conclusions

In conclusion, the increase in blood NAD concentrations after NMN supplementation has a high interindividual variability and a higher NAD increase is associated with better functional outcomes and quality of life. Therefore, blood NAD concentrations should be measured during NMN supplementation. Further studies on personalized NAD precursor supplements and optimal blood NAD concentration are warranted.

## Ethical approval

The ethics committee is duly registered with Drugs Controller General of India (DCGI) via number - ECIV45/Indt/MII/2013/RR-19. The trial was monitored by ProRelix Services LLP, a clinical research organization (CRO), Pune, India.

## Trial registration

NCT04823260 in ClinicalTrials.gov, and CTRI/2021/03/032421 in Clinical Trial Registry – India.

## Informed consent

Written informed consent was obtained from all participants.

## Author contributions

Conceptualization: A.H.K., W.W, L.Y., and A.B.M. Methodology: A.H.K. and W.W. Resources: L.Y., R.T., Z.L., A.V., S.P., S.T., N.A., G.A., V.K., and A.B.M. Statistics: W.W. Supervision: A.B.M. Writing: all authors.

## Materials and correspondence

Material request or correspondence to Andrea B. Maier

## Data availability

Data available on request from the authors.

